# Optimal adherence thresholds for oral anticoagulants in patients with atrial fibrillation using machine learning and population administrative data

**DOI:** 10.1101/2025.06.06.25329171

**Authors:** Abdollah Safari, Hamed Helisaz, Mina Tadrous, Marc W. Deyell, Jason G. Andrade, Shahrzad Salmasi, Adenike Adelakun, Kristian B. Filion, Mary A. De Vera, Peter Loewen

**Affiliations:** School of Mathematics, Statistics, and Computer Science, College of Science, University of Tehran, Iran; Department of Statistics, Faculty of Science, University of British Columbia, Vancouver, Canada; Leslie Dan Faculty of Pharmacy, University of Toronto. Toronto, Canada; Women’s College Hospital, Toronto, Canada; Division of Cardiology, Faculty of Medicine, University of British Columbia, Vancouver, Canada; UBC Center for Cardiovascular Innovation, Vancouver, Canada; Centre for Health Evaluation & Outcome Sciences, Providence Health Care Research Institute, Canada; Atrial Fibrillation Clinic, Electrodiagnostics, and Cardiac Electrophysiology, Vancouver General Hospital, Canada; Montreal Heart Institute, Université de Montréal, Montréal, Canada; Faculty of Pharmaceutical Sciences, University of British Columbia, Vancouver, Canada; Collaboration for Outcomes Research and Evaluation, Faculty of Pharmaceutical Sciences, University of British Columbia, Canada; Centre for Clinical Epidemiology, Lady Davis Institute, Jewish General Hospital, McGill University, Montreal, Canada

**Keywords:** atrial fibrillation, medication adherence, oral anticoagulants, stroke, major bleeding

## Abstract

**Background:** Adherence to oral anticoagulants (OACs) for atrial fibrillation (AF) stroke prevention is traditionally defined as taking 80% of doses as prescribed, but this threshold has not been validated in terms of clinical outcomes.

**Objectives:** We sought to determine the OAC adherence thresholds maximally associated with risk of clinical events in patients with AF. We also sought to compare these thresholds to the conventional 80% threshold.

**Methods:** This was a cohort study using retrospective data of patients newly diagnosed with AF who were new users of OACs from 1996 to 2019 from population-based administrative data from British Columbia, Canada. We built Cox proportional hazard models with OAC adherence, measured as proportion of days covered (PDC), as the main exposure captured during 90 days before outcome events or end of follow-up and applied LASSO estimated coefficient paths to identify candidate thresholds, followed by steps to identify which were truly optimal in terms of outcomes.

**Results:** A total of 44,172 patients were included. For all outcomes, VKA optimal PDC thresholds were between 0.85 and 0.95 and the DOAC optimal threshold was 0.9. Outcome hazard reduction for adherence above vs. below the optimal threshold was greater for DOAC than VKA. For DOAC, PDC ≥0.9 yielded risk reductions of 19 to 52% compared to being below it for stroke outcomes, while for these were not significant for VKA for most outcomes. The conventional PDC 0.8 threshold had inferior model fit and clinical relevance compared to the optimal thresholds identified.

**Conclusions:** Optimal adherence thresholds are higher than conventionally assumed and, especially with DOACs, are strongly associated with clinical outcomes. Our results highlight the need to consider updating the definition of nonadherence to PDC <0.9 for DOACs, and to PDC<0.9 or <0.95 for VKA, depending on the outcome.

## INTRODUCTION

Nonadherence to oral anticoagulants (OACs) in patients with atrial fibrillation (AF) is common^1,2^ and associated with substantially worse health outcomes, particularly stroke and death.^3–10^ Medication adherence is typically measured as a continuous variable ranging from 0 (completely nonadherent) to 1 (completely adherent) using the proportion of days covered (PDC) or medication possession ratio (MPR).^11^ Given the difficulty of achieving complete adherence in practice,^12–14^ dichotomizing continuous variables using thresholds is valuable to define “how much is good enough?” and for classifying patients in quality-of-care measurement programs,^12^ identifying patients in need of adherence-enhancing interventions or a different medication, and for medication adherence research.^13,14^ However, where thresholds are used, they must predict clinical outcomes for the disease involved.^14^ Otherwise, they may be insensitive to patients in need of interventions, needlessly penalize providers, divert resources to unnecessary adherence-improvement programs, or provide false assurance of the effectiveness of a patient’s medication regimen. In the case of OAC adherence in patients with AF, a miscalibrated adherence threshold could result in avoidable strokes and deaths.

A threshold of 80% is widely used to classify OAC adherence in patients with AF.^2,9,15–18^ However, few studies have attempted to identify and evaluate the clinical significance of thresholds for OACs in AF,^19–21^ and none have attempted to find thresholds associated with the highest model performance, predictive accuracy, or effect on clinical outcomes. Hence, clinically significant, evidence-based (i.e. “optimal”) adherence thresholds for OACs in the treatment of AF remain unknown. Furthermore, the optimal thresholds may vary by outcome, drug or drug class, and more than one optimal threshold may exist for any of these scenarios.^22^

Our primary objective was to identify outcome-specific OAC adherence thresholds which are associated with the risk of clinical events (stroke or systemic embolism (SSE), SSE or transient ischemic attack (TIA) or death, SSE or TIA, ischemic stroke, death, cardiovascular death, major bleeding) and are therefore valid for discriminating adherent from nonadherent patients with AF. Our secondary objectives were to compare these new thresholds to the conventional 80% threshold and compare the performance of several models representing the relationship between PDC and outcome risk.

## METHODS

### Study design and setting

This was a cohort study using retrospective de-identified patient records from administrative data (Population Data BC) for the entire population of the province of British Columbia, Canada (∼5 million residents) from 1996 to 2019. Datasets included outpatient billing, details of hospitalizations, participant demographics, date and primary cause of death, and data for all prescription medications dispensed outside of the hospital (PharmaNet). Person-level linkage used a study-specific unique identifier created by the data provider and linkage was performed before the data was released to the investigators. Details of the data sources, which are used extensively for patient outcomes research^23^ are available at https://www.popdata.bc.ca/data.

The study was approved by the University of British Columbia Clinical Research Ethics Board (H22-02598). Study reporting follows the Strengthening the Reporting of Observational Studies in Epidemiology (STROBE) and Reporting of Studies Conducted Using Observational Routinely Collected Health Data Statement for Pharmacoepidemiology (RECORD-PE) extension guides.^24–26^

### Study population

Details of the study cohort definition have been published previously.^10^ Briefly, we developed an incident cohort of adults with non-valvular AF via validated algorithms using outpatient billing codes and hospitalization codes (Supplemental Table 1). Next, we limited the cohort to incident OAC users (warfarin, dabigatran, apixaban, rivaroxaban, or edoxaban) excluding those with a non-AF indication for anticoagulation using previously published methods^10^ (Supplemental Figure 1). Follow-up began on the date of the second OAC prescription fill after the AF diagnosis date, since measurement of the exposure variable, proportion of days covered (PDC), requires ≥1 prescription refill. Cohort entry (“index date”) occurred on the date of the second OAC prescription fill. Because index date and cohort entry were the same and outcomes and exposures were time-dependent, risk of bias due to immortal time was minimized.^27^ Potential bias from patients having events between diagnosis date (which was before cohort entry) and index date were minimized by including such events as a covariate in study models. Follow-up ended at the time of first study event, gap in plan enrollment of >30 days, or on the date of the last available data, whichever came first. (Figure 1).

**Figure 1:**
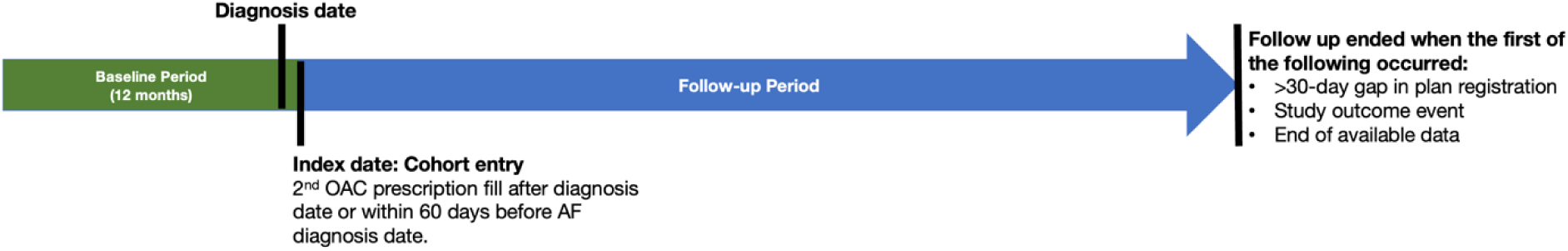
Study design

### Exposures

The primary exposure of interest was PDC during 90 days prior to events calculated using standard methods.^28–30^ To address variable VKA dosing, we used the validated Random Effects Warfarin Days’ Supply (REWarDS) method^31^ to estimate days’ supply, as we have previously.^10,32^ REWarDS is more accurate than other methods for estimating PDC from VKA prescription records.^31^ At this stage we did not exclude patients who appeared to discontinue OAC therapy permanently. To enable analyses of association between OAC drug classes and the study outcomes, we created an additional variable for OAC at time of the event using a novel scheme described in Supplemental Methods 1. Briefly, to study the effect of OAC adherence and OAC drug class separately, we included their corresponding information in three separate variables: PDC, OAC drug class, and individual OAC drug. The former measured patients’ adherence to any prescribed OAC medication calculated during 90-day window prior to the event date while accounting for stockpiling, or end of follow-up, and the others captured the class or drug temporally associated with the event. Patients whose OAC supply ran out during the 90 days prior to their event (i.e., were recent users but not active users at the time of their event) were classified as inactive.

### Outcomes

The primary outcomes were the composite endpoint of SSE, TIA, or death, and SSE as an individual endpoint. Secondary outcomes were a composite endpoint of SSE or TIA, and the individual endpoints of ischemic stroke, all-cause death, cardiovascular death, and major bleeding. All outcomes were measured as time to first event and were based on the first or second most responsible cause of hospitalization. Validated algorithms were used to ascertain all outcomes (Supplemental Table 2).

### Covariates

Patient characteristic covariates were measured during 12 months prior to cohort entry and included age, sex, SSE risk score (CHA_2_DS_2_-VASc; Congestive heart failure, Hypertension, Age ≥75, Diabetes, prior Stroke or systemic embolism, Vascular disease, Age 65-74, Sex [female]),^33^ major bleeding risk score (modified HAS-BLED; Hypertension, Abnormal renal/liver function, Stroke, Bleeding, Labile INR, Elderly, and Drugs/alcohol),^34,35^ comorbidities (individual components of CHA_2_DS_2_-VASc and HAS-BLED scores), socioeconomic status (SES), neighbourhood income quintile, number of medical services plan billings, number of hospitalizations, time from AF diagnosis to OAC start, Charlson and Elixhauser comorbidity indices,^36,37^ and polypharmacy (5 or more concurrent medications, not including OAC).^38^ Following convention, CHA_2_DS_2_-VASc and HAS-BLED scores were dichotomized at ≥2. Due to their highly skewed distributions, number of hospitalizations during follow-up was grouped into four categories (0, 1, 2, and ≥3), and time to OAC initiation was grouped into three categories (<3, 3-9, >9 months). Multivariable imputation by chained equations was used to impute missing values for SES, the only covariate with missing values.^39^

### Statistical analyses

To evaluate the association between OAC adherence and risk of clinical outcomes, multivariable Cox proportional hazard models were constructed that incorporated PDC as the main exposure, and the other study covariates (described above). For all analyses, PDC was the value during the 90-day window ending with their outcome event or end of follow-up for those without an outcome event. As such, results were based on adherence in immediate temporal proximity to the outcome event.

### Optimal thresholds using machine learning

Thresholds, also called cut-off values or cut points, represent specific points above which patients exhibit an elevated or reduced risk of an outcome. We defined ‘optimal thresholds’ as those PDC values at which the disparity in outcome risk is most pronounced between patients with PDC values below and above them. Optimal thresholds were identified via a 3-stage process.

In Stage 1, continuous PDC was transformed into binary variables at 5% intervals, and the biniLasso method, a specially designed LASSO (Least Absolute Shrinkage and Selection Operator) regularized Cox model, was applied to identify candidate thresholds, focusing on the first one or two for simplicity and interpretability.^40^ The LASSO regularization helps to pinpoint the most important regions across PDC spectrum (corresponding to the top PDC thresholds) where its association with the outcome exhibited the most pronounced changes according to the Cox partial likelihood.^41^ These thresholds were used to construct either a binary (low/high) or three-level (low/middle/high) categorical PDC, resulting in a “1-threshold” or “2-threshold” Cox model, respectively. The final model for Stage 2 was selected based on Akaike’s Information Criterion (AIC). Full methodologic detail is provided in a separate paper.^40^

In Stage 2, the viability of candidate thresholds was assessed by evaluating the number of outcome events within each adherence category created by the threshold. To maintain statistical rigor, thresholds were considered viable only if there were at least 5 events within the exposure strata.^42^ If this criterion was not met for a given threshold, we defaulted to the model with the next-lowest number of thresholds. If the 1-threshold model also failed to meet the EPV criterion, we substituted the second threshold from the 2-threshold model as an alternative 1-threshold model and subjected it to the same criterion.

In Stage 3, we evaluated the effects of the thresholds on outcome hazard, with the principle that a threshold should have a statistically significant association with outcome hazard to be considered optimal. This was done by conducting contrast tests on the outputs of the fitted Cox models with PDC strata classified as low/middle/high or low/high, depending on the number of thresholds in the model carried forward from previous stages. Two versions of hazard ratio (HR) were generated for each threshold: The PDC-HR represented the relative hazard of event for patients actively taking OAC at time of event (based on having supply available) who were above the threshold compared to patients with identical covariate characteristics who were below the threshold. The OAC-HR represented the relative hazard of event when PDC increased from low to high for actively OAC-taking patients compared to inactive OAC users with identical covariate characteristics. This captured the combined effects of being an active OAC user (compared to inactive) and of PDC. Thresholds with a nonsignificant PDC-HR and OAC-HR for all OAC drug classes were abandoned, and the model with one fewer threshold was selected and subjected to the same criterion. If no threshold met the criteria in the 1-threshold model, identification of an optimal threshold was abandoned for that outcome. Thresholds persisting through all three stages were retained and reported as the “optimal threshold(s)” for each outcome.

### Comparing optimal thresholds to the conventional threshold (PDC 0.8)

To compare our derived optimal thresholds to the conventional threshold (PDC 0.8) we estimated the 95% confidence interval (CI) around each optimal threshold using 1000 bootstrapped samples randomly drawn from the full cohort. If this interval excluded 0.8, the conventional threshold was considered different than the one we derived.

To further evaluate whether our newly derived optimal thresholds were superior to the conventional threshold, we compared the relative fidelity of the 1-threshold, 2-threshold, and conventional threshold (PDC 0.8) models by using 10,000-iteration bootstrap resampling from the entire cohort to generate 95% confidence intervals for AIC. Then we did pairwise paired-sample t-tests to compare these AIC values for each of the 3 models, for each outcome, with p-value adjustment for multiple testing using Holm’s method.^43^

### Model performance comparison

To evaluate the trade-off involved in using simpler (e.g., 1-threshold categorical) vs. more complex models (e.g., continuous b-spline), we compared the performance of four different models: continuous b-spline, linear, and the previously described 1- and 2- threshold categorical models. The continuous b-spline model was an additive Cox proportional hazards model implemented through a Cox-based Generalized Additive Model (CGAM),^44^ a regression algorithm that employs smoothing splines, for each outcome. The linear model was a standard Cox model with PDC as a continuous exposure and linearly associated with each outcome. For each, we generated 95% confidence intervals for its AIC using 10,000 bootstrap resampling, then for each outcome, all AICs were compared to each other with paired-sample t-tests with Holm p-value adjustment.^43^

### Sensitivity analysis

We tested our approach of including inactive users in study models by performing our analyses on the primary outcomes with those patients excluded. All analysis was conducted using R v4.0.5 (R Core Team, Vienna, Austria, 2021) and RStudio v1.3.1039 (RStudio Team, Boston, US, 2020).

## RESULTS

### Participants

The study cohort included 44,172 patients with mean age 70.6 years (SD 11.4), 44% of whom were female. Of these, 12,928 were classified as VKA users at the time of their primary outcome event, 16,688 as DOAC users, and 14,556 as inactive. Median follow-up time was 6.8 years [interquartile range (IQR): 4.1, 10.9]. Other participant characteristics are shown in Table 1. A study flow diagram is shown in Supplemental Figure 1. Event frequencies are shown in Supplemental Table 3.

**Table 1:**
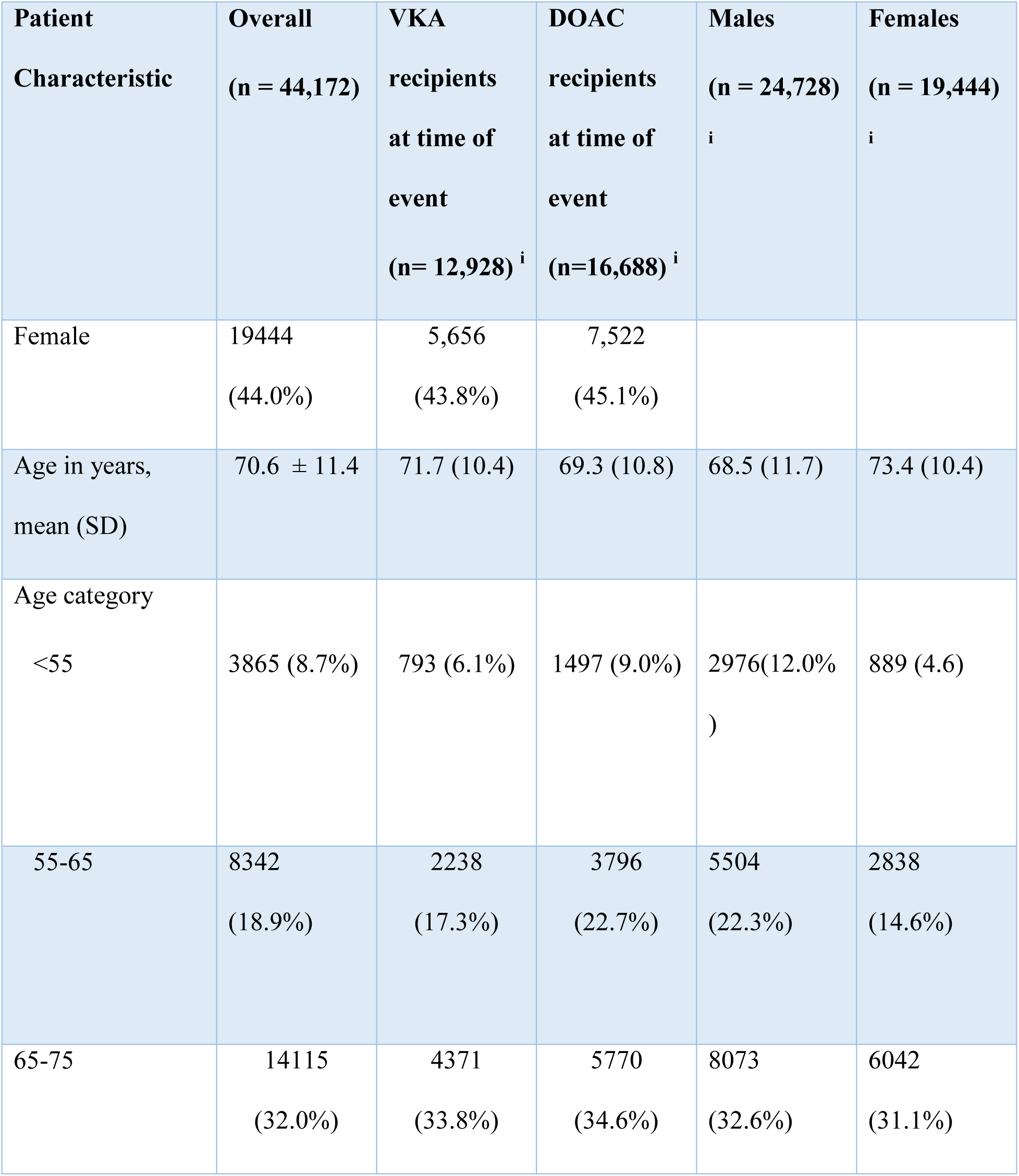
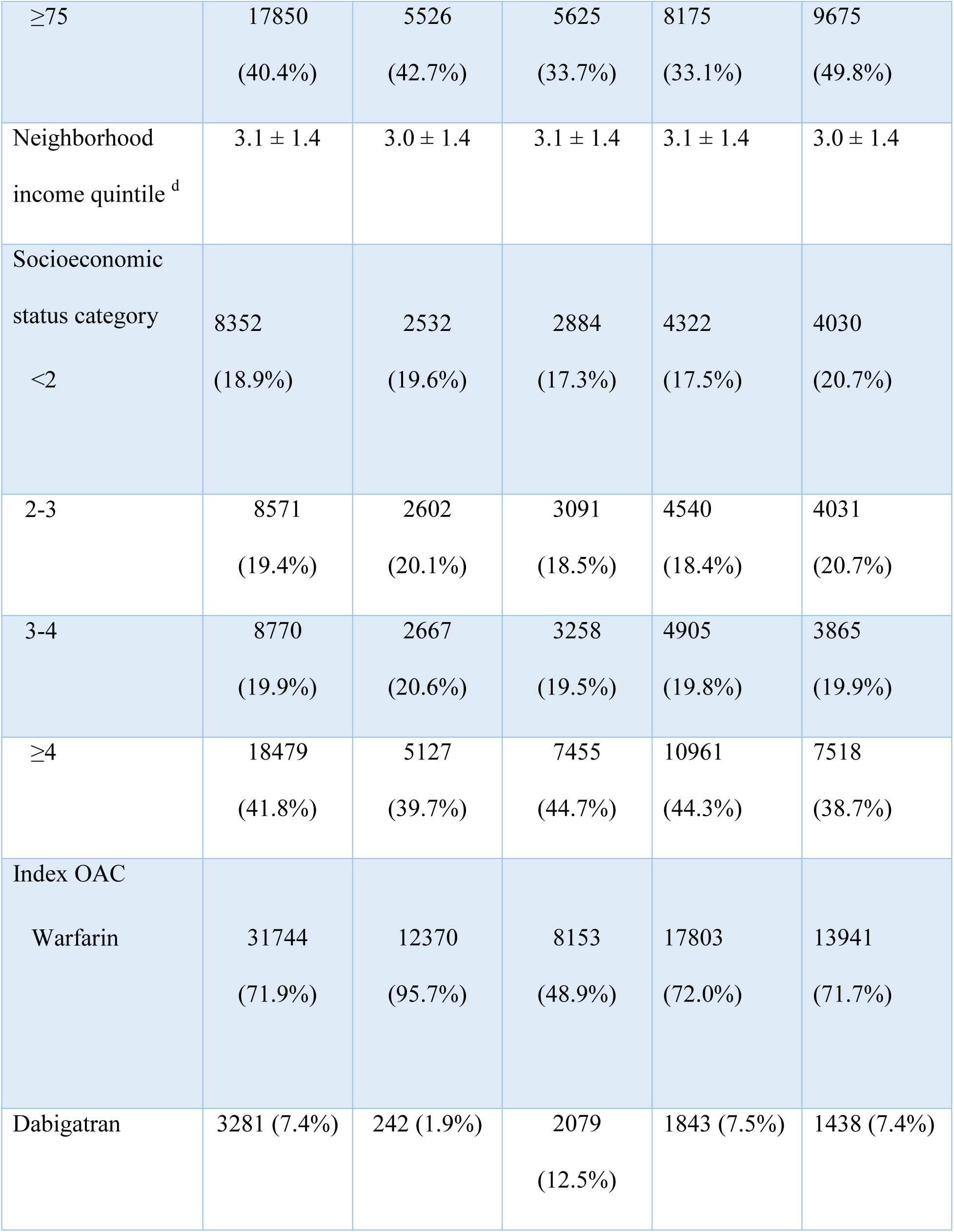
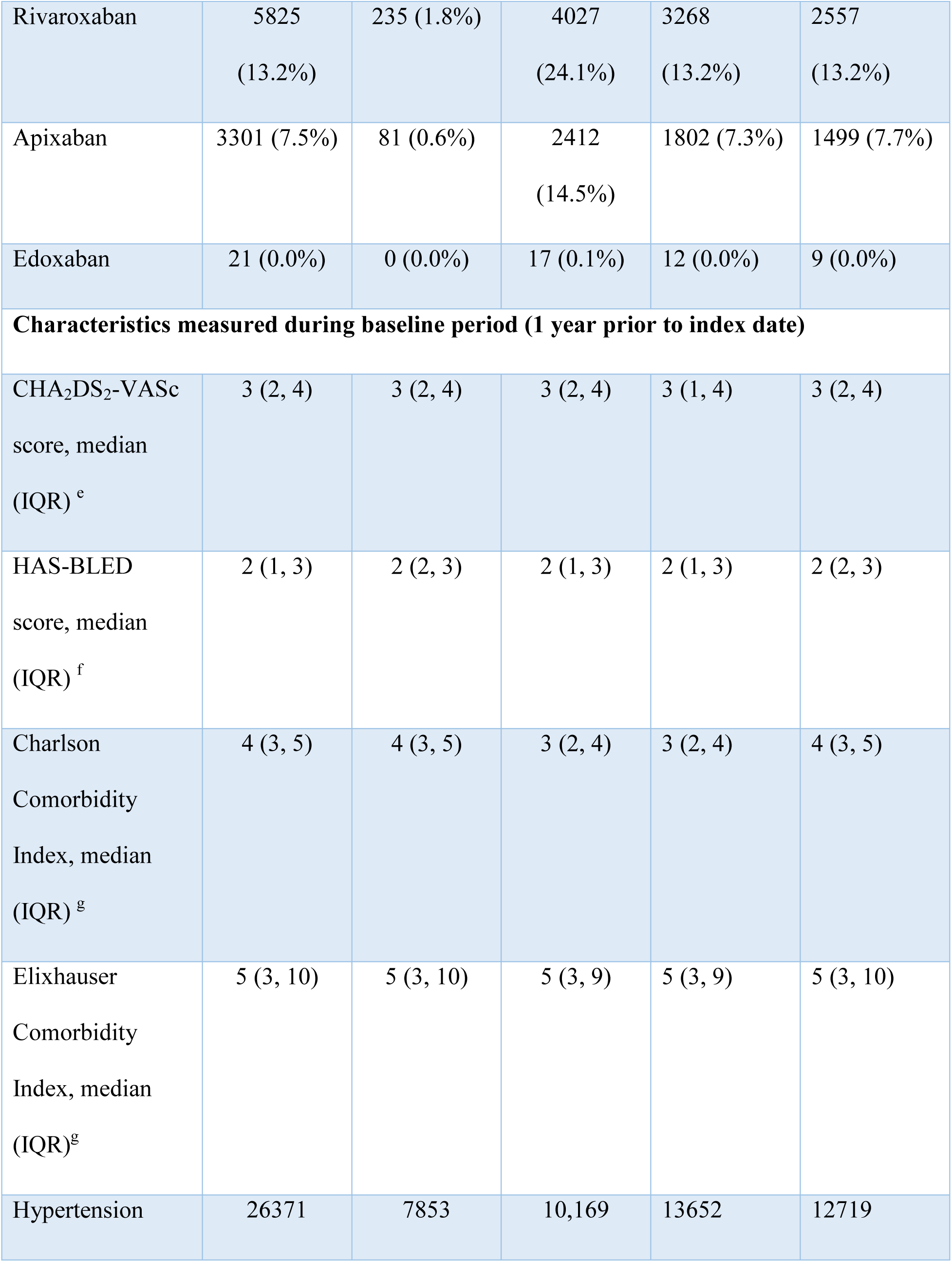
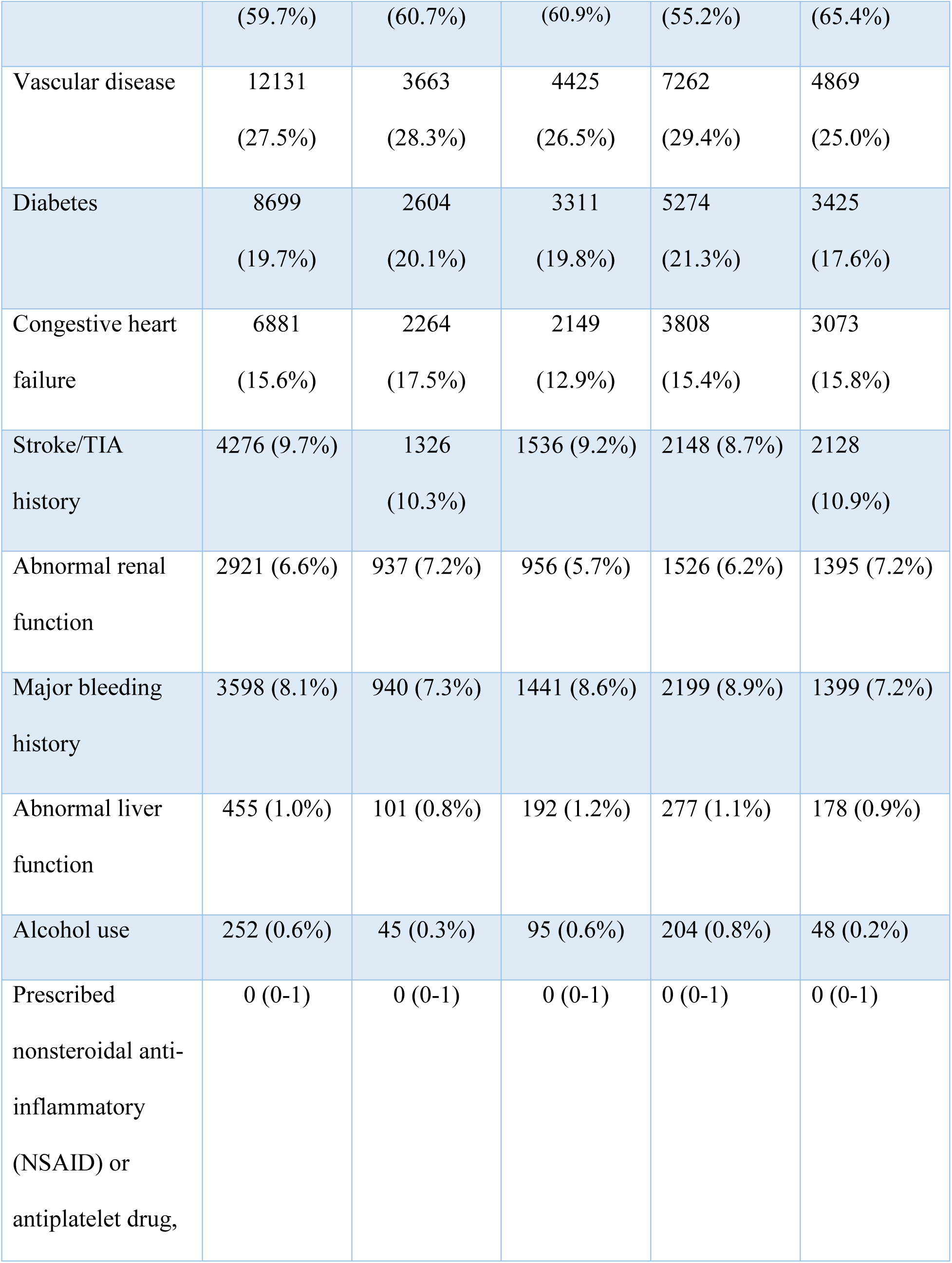
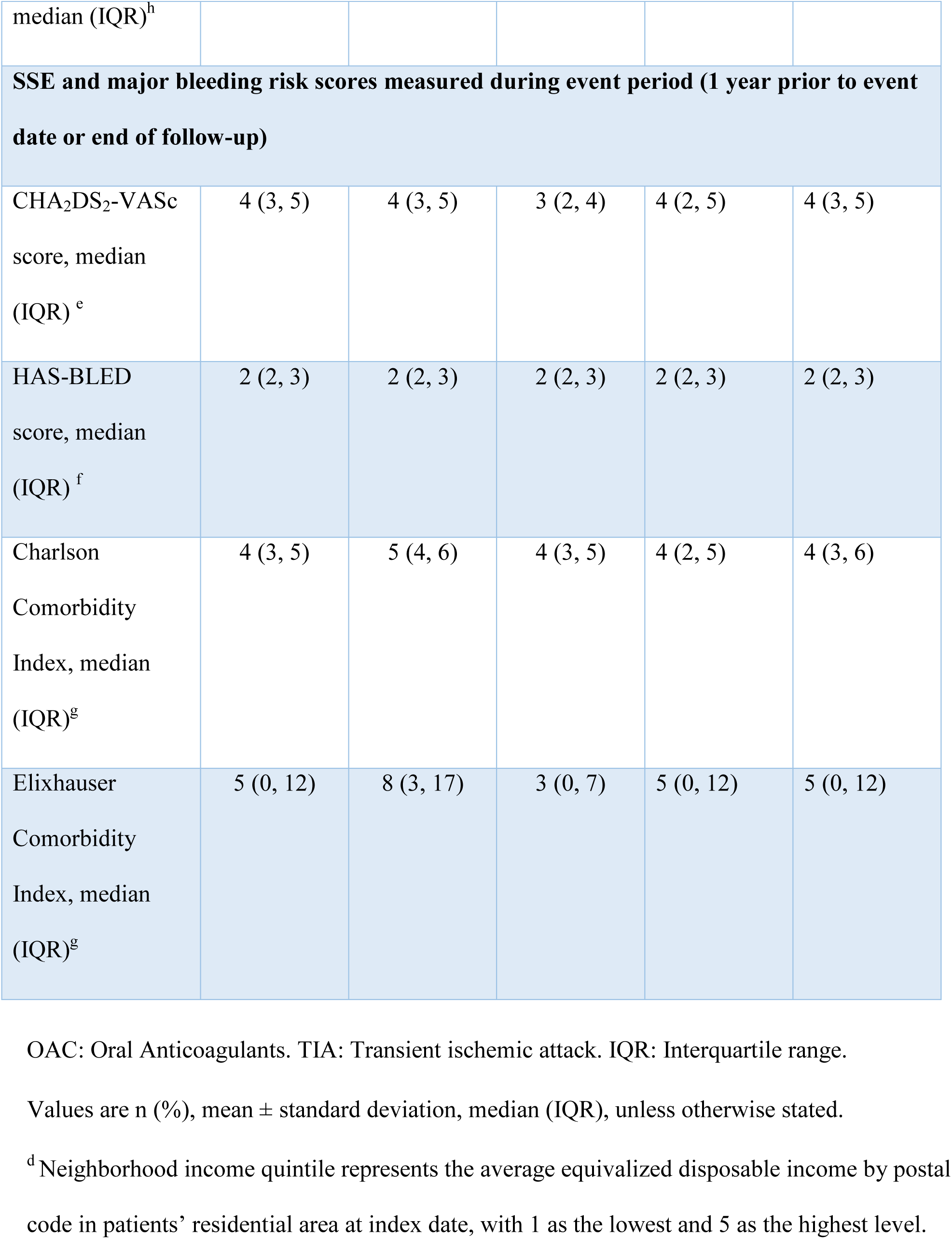
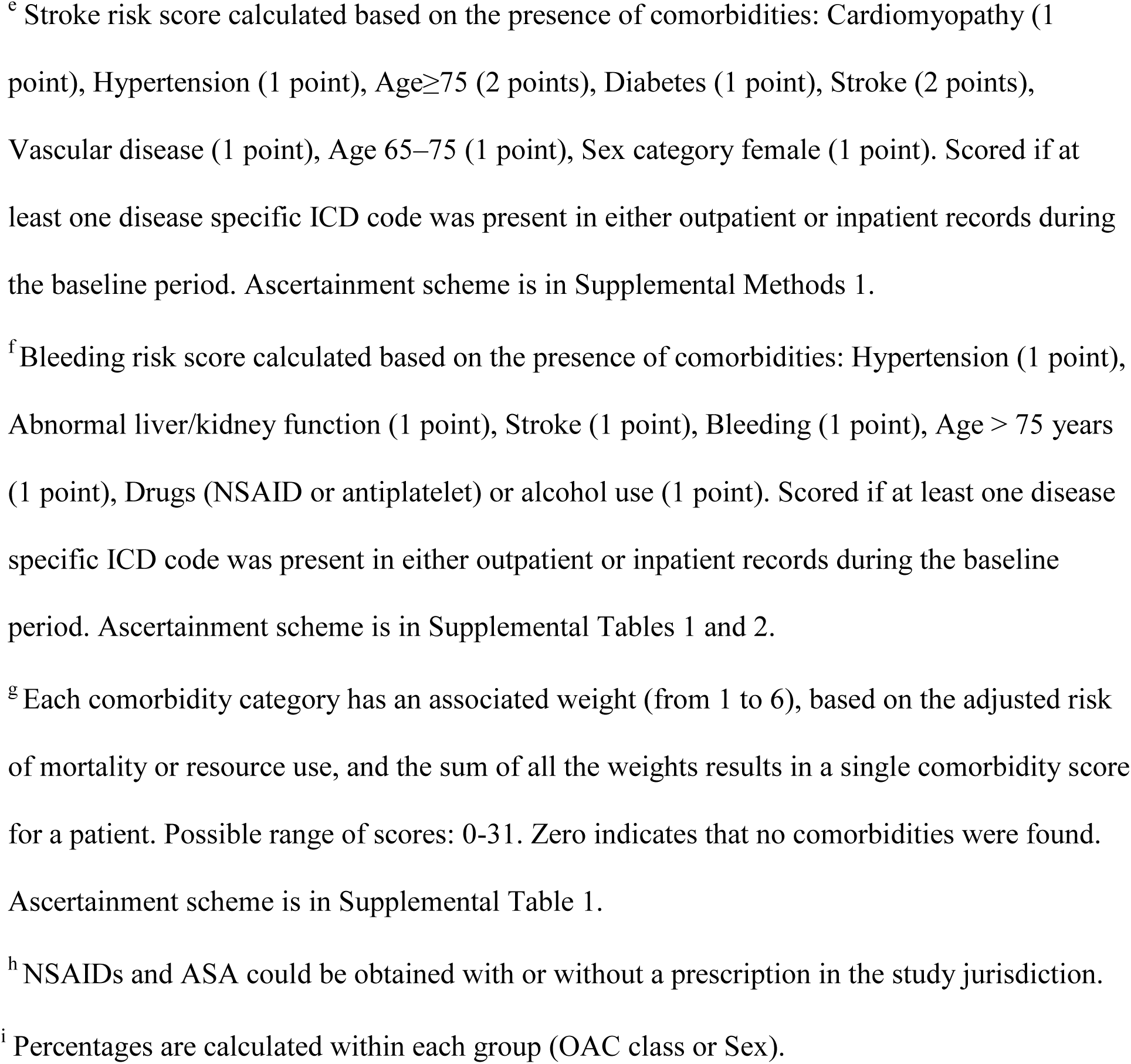
Characteristics of study patients at index date.

### Optimal adherence thresholds

Optimal thresholds and their associations with outcome hazards from categorical modelling are shown in Table 2 and Figures 2 and 3. For DOACs, the single optimal threshold for all outcomes was PDC 0.90. PDC-HRs for outcomes involving stroke ranged from 0.81 (95%CI 0.72-0.91) for SSE, TIA, or death to HR 0.48 (95%CI 0.37-0.63) for ischemic stroke, strongly implying the clinical importance of adherence above the optimal threshold compared to below it. PDC-HRs for death outcomes were 0.91 (0.80-1.04) for all-cause death and 0.88 (0.72-1.08) for cardiovascular death. The OAC-HRs showed larger effect sizes (33-69% risk reductions) since they include the additional effect of being an active OAC user at time of event vs. being inactive. Figures 2 and 3 depict the negative association between PDC and outcome hazard for all effectiveness outcomes, which is particularly marked for DOAC. The LASSO coefficient paths used in stage 1 are shown in Supplemental Results 1.

**Figure 2:**
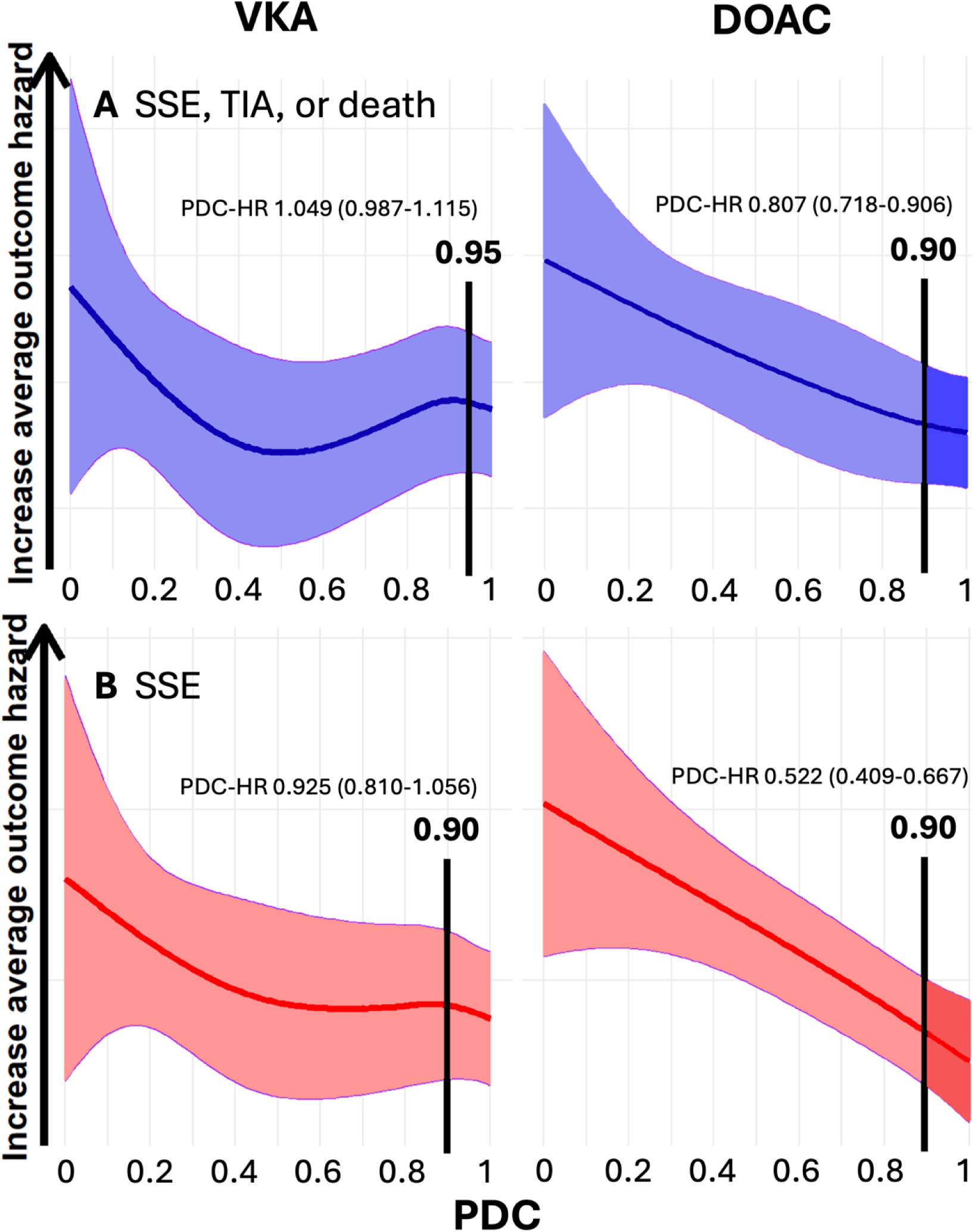
Continuous spine plot of **A.** SSE, TIA, or death and **B.** SSE hazard over the PDC spectrum with superimposed optimal PDC thresholds and PDC hazard ratio for identical patients above vs. below the optimal threshold.

**Figure 3:**
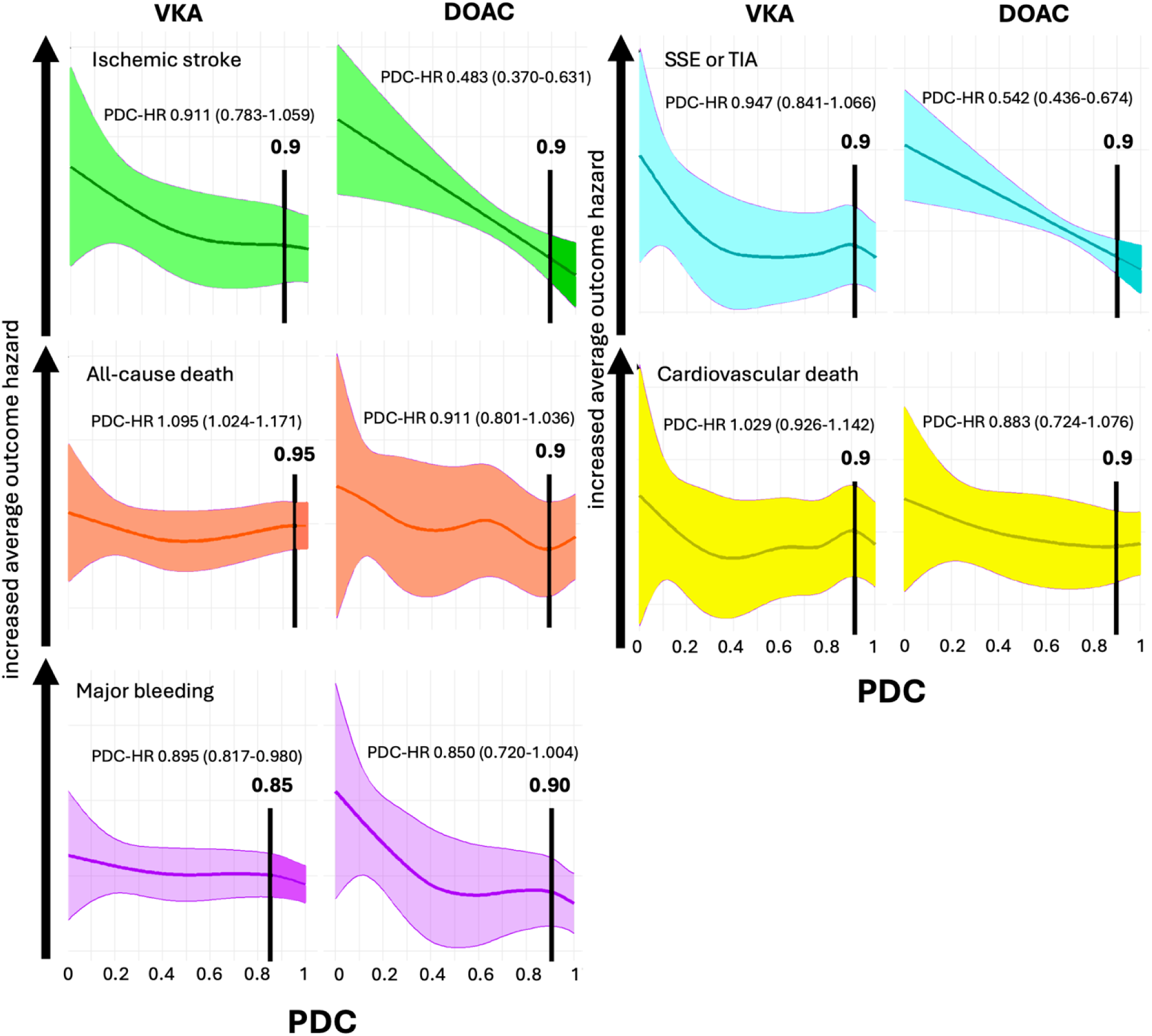
Secondary outcomes; Continuous spine plot of outcome hazard over the PDC spectrum with superimposed optimal PDC thresholds and PDC hazard ratio for identical patients above vs. below the optimal threshold.

**Table 2:** Optimal PDC thresholds and magnitude of effect on outcome hazard by drug class.

|  | VKA |  |  | DOAC |  |  | Notes |
| --- | --- | --- | --- | --- | --- | --- | --- |
|  | Optimal threshold<br>(95% CI)* | PDC-HR<br>HR<br>(95% CI) | OAC-HR<br>HR<br>(95% CI) | Optimal threshold<br>(95% CI)* | PDC-HR<br>HR<br>(95% CI) | OAC-HR<br>HR<br>(95% CI) |  |
| **SEE, TIA, or death** | 0.95<br>(0.948-0.952) | 1.049<br>(0.987-1.115) | 0.739<br>(0.674-0.811) | 0.90<br>(0.897-0.903) | 0.807<br>(0.718-0.906) | 0.569<br>(0.497-0.651) | 2-threshold model (VKA: 0.9, 0.95; DOAC: 0.8, 0.9) had lowest AIC of categorical models, but was rejected at stage 3 for non-significant HRs for DOAC and VKA. Hence, 1-threshold model selected. |
| **SSE** | 0.90<br>(0.896-0.904) | 0.925<br>(0.810-1.056) | 0.584<br>(0.488-0.699) | 0.90<br>(0.895-0.905) | 0.522<br>(0.409-0.667) | 0.330<br>(0.251-0.433) | 2-threshold model (VKA: 0.85, 0.9; DOAC 0.8, 0.9) had lowest AIC of categorical models, but was rejected at stage 2 for insufficient outcome events in some strata. |
| **SSE or TIA** | 0.90<br>(0.897-0.903) | 0.947<br>(0.841-1.066) | 0.599<br>(0.509-0.704) | 0.90<br>(0.895-0.905) | 0.542<br>(0.436-0.674) | 0.343<br>(0.269-0.437) | 2-threshold model (VKA: 0.85, 0.9; DOAC 0.8, 0.9) had lowest AIC of categorical models, but was rejected at stage 2 for insufficient outcome events in some strata. |
| **Ischemic stroke** | 0.90<br>(0.893-0.907) | 0.911<br>(0.783-1.059) | 0.643<br>(0.525-0.788) | 0.90<br>(0.895-0.905) | 0.483<br>(0.370-0.631) | 0.341<br>(0.253-0.460) | 2-threshold model (VKA: 0.9, 0.95; DOAC 0.8, 0.9) had lowest AIC of categorical models, but was within 1 point of 1-threshold model AIC so 1-threshold model was pursued. |
| **All-cause death** | 0.95<br>(0.948-0.951) | 1.095<br>(1.024-1.171) | 0.794<br>(0.718-0.878) | 0.90<br>(0.896-0.904) | 0.911<br>(0.801-1.036) | 0.661<br>(0.569-0.767) | 2-threshold model (VKA: 0.9, 0.95; DOAC 0.75, 0.9) had lowest AIC of categorical models, but was rejected at stage 2 for insufficient outcome events in some strata. |
| **Cardiovascular death** | 0.90<br>(0.897-0.903) | 1.029<br>(0.926-1.142) | 0.692<br>(0.592-0.810) | 0.90<br>(0.897-0.903) | 0.883<br>(0.724-1.076) | 0.594<br>(0.473-0.747) | 2-threshold model (VKA: 0.9, 0.95; DOAC 0.9, 0.95) had lowest AIC of categorical models, but was rejected at stage 2 for insufficient outcome events in some strata. |
| **Major bleeding** | 0.85<br>(0.846-0.853) | 0.895<br>(0.817-0.980) | 0.324<br>(0.273-0.383) | 0.90<br>(0.885-0.915) | 0.850<br>(0.720-1.004) | 0.307<br>(0.247-0.283) | 2-threshold model (VKA: 0.85, 0.8; DOAC 0.35, 0.9) had lowest AIC of categorical models, but was rejected at stage 2 for insufficient outcome events in some strata. |

AIC: Akaike Information Criterion
NS: nonsignificant
PDC-HR: hazard of event for actively VKA-taking patients with identical covariate values who are above vs. below PDC optimal threshold
OAC-HR: hazard of event when PDC increases from low to high for actively VKA-taking patients compared to inactive OAC users with identical covariate characteristics
\* confidence intervals were derived from 1000 iteration bootstrap sampling

For VKA, across effectiveness outcomes the single optimal threshold was either 0.90 or 0.95. For all outcomes except all-cause death, PDC-HR magnitudes were smaller than for DOACs and not statistically significant, implying a relatively flat relationship between PDC and outcome hazard across the PDC spectrum. Figures 2 and 3 support this interpretation. On the other hand, the OAC-HR effect sizes were large (21-67% risk reductions) and significant for all effectiveness outcomes with VKA, showing that, for the same increase in PDC from low to high, being an active VKA user at time of event significantly reduced outcome hazard compared to being inactive.

The results were slightly different for major bleeding. The single VKA optimal threshold was PDC 0.85 and the PDC-HR was statistically significant [HR 0.90 (95%CI 0.82-0.98)]. For DOAC the optimal threshold was 0.90 and its PDC-HR was borderline significant [HR 0.85 (0.72-1.004)]. The OAC-HRs were large and significant for VKA and DOAC.

### Optimal adherence thresholds vs. conventional PDC 0.8 threshold

Results of the bootstrapped AIC analyses comparing the double and single optimal threshold models to a PDC 0.8 threshold model for each outcome are shown in Table 3. For all outcomes, both the single and double optimal thresholds models were superior to the conventional PDC 0.8 threshold model.

**Table 3:** Comparison of derived threshold models to conventional (PDC 0.8) threshold models using AIC.

| **Outcome** | **AIC difference (95%CI) \*** |  |
| --- | --- | --- |
|  | **1-threshold vs. conventional (PDC 0.8) threshold model** | **2-threshold vs. conventional (PDC 0.8) threshold model** |
| SEE, TIA, or death | -25.8 (-26.1 to -25.5) <sup>Δ</sup> | -117.0 (-117.5 to -116.6) <sup>Δ</sup> |
| SSE | -13.6 (-13.9 to -13.2) <sup>Δ</sup> | -23.4 (-23.7 to -23.1) <sup>Δ</sup> |
| SSE or TIA | -5.3 (-5.6 to -4.9) <sup>Δ</sup> | -18.1 (-18.5 to -17.8) <sup>Δ</sup> |
| Ischemic stroke | -2.4 (-2.6 to -2.1) <sup>Δ</sup> | -5.4 (-5.6 to -5.2) <sup>Δ</sup> |
| All-cause death | -30.8 (-31.1 to -30.6) <sup>Δ</sup> | -45.1 (-45.4 to -44.8) <sup>Δ</sup> |
| Cardiovascular death | -23.1 (-23.2 to -22.9) <sup>Δ</sup> | -39.2 (-39.4 to -38.9) <sup>Δ</sup> |
| Major bleeding | -6.4 (-6.5 to -6.3) <sup>Δ</sup> | -24.9 (-25.1 to -24.7) <sup>Δ</sup> |

AIC: Akaike Information Criterion
\* Derived from 10000 bootstrap resampling
<sup>Δ</sup> p<0.0001 from paired-sample t-test with Holm p-value adjustment

As shown in Table 2, all optimal thresholds we identified were statistically significantly different from 0.80 based on their bootstrap sampled 95% confidence intervals. A threshold of 0.8 emerged in some 2-threshold models for DOAC, which were obviously not different from the conventional threshold, but these thresholds were rejected for the reasons described in Table 2. Rejected thresholds close to 0.8 emerged for some outcomes (e.g. VKA 0.85 for SSE, SSE or TIA, major bleeding; DOAC 0.75 for all-cause death) but in each case they were statistically significantly different from 0.8 and rejected due to insufficient outcome events in some strata. See Supplemental Results 2 for detailed model outputs.

### Model Performance Comparisons

Model performance corresponded to model complexity, with continuous b-spline (highest complexity) showing highest performance and 1-threshold categorical (lowest complexity) demonstrating lowest performance, with all AIC differences being highly statistically significant (Table 4). This was consistent with positive nonlinearity tests for the CGAM models (Supplemental Table 4).

**Table 4:** Akaike Information Criterion (AIC) to compare the fitted Cox model performance. Lowest AIC (best model fit) is shown in bold text.

| Outcome | Continuous b-spline model | Linear model | 2-threshold categorical model | 1-threshold categorical model |
| --- | --- | --- | --- | --- |
| SEE, TIA, or death | **327056.1** | 327264.4 | 327351.6 | 327439.7 |
| SSE | **64015.9** | 64105.2 | 64113.9 | 64120.7 |
| SSE or TIA | **79784.7** | 79927.6 | 79948.5 | 79958.4 |
| Ischemic stroke | **50788.8** | 50866.7 | 50880.6 | 50880.6 |
| All-cause death | **300769.8** | 301000.1 | 301164.9 | 301238.3 |
| Cardiovascular death | **127238.5** | 127333.1 | 127401.8 | 127414.9 |
| Major bleeding | **121589.2** | 121631.8 | 121816.0 | 121812.7 |

AIC: Akaike Information Criterion
95% confidence intervals were computed for each AIC using 10,000 bootstrap resampling, then for each outcome, all AICs were compared to each other with paired-sample t-tests with Holm p-value adjustment. All comparisons were significantly different with $p < 0.0001$

### Sensitivity analysis

After excluding inactive patients (n=14,323) and applying the same three-stage approach to identifying optimal thresholds, a single optimal threshold was identified for both primary outcomes, which were 0.9 for DOAC and 0.95 for VKA. Results of this sensitivity analysis are in Supplemental Results 3.

## DISCUSSION

In this large population-based observational study using routinely collected health administrative data, the optimal adherence thresholds for all outcomes for both VKA and DOAC were robustly confirmed to be higher than the conventional threshold of 80%. Furthermore, the clinical significance of thresholds for most outcomes was much greater for DOAC than VKA, based on hazard reduction. In fact, for nearly all outcomes, being above or below the threshold was not associated with significantly different outcome hazard for VKA users, whereas for DOAC, being above PDC 90% yielded risk reductions of 19 to 52% compared to being below it for outcomes involving stroke.

The threshold of 80% for medication adherence was first used in the late 1970s when it was arbitrarily applied to antihypertensive medications^13^ and was adopted for other diseases and treatments with almost no validation of its clinical significance.^1,45,46^ A 2022 scoping review found that of 76 studies using adherence thresholds, only 3 tried to identify appropriate thresholds, 80% used the conventional PDC 0.8, and few (18%) cited evidence of the clinical relevance of the chosen threshold to justify it.^28^ In the few chronic conditions where the 80% threshold has been challenged, it has shown poor predictive accuracy for clinical outcomes.^14,22,47–53^ In AF, use of the 80% threshold is widespread in OAC studies and programs despite minimal evaluation of its clinical relevance.^19–21,54^ Previous studies of OAC thresholds in AF did not attempt to identify optimal thresholds, instead comparing categories of average PDC over follow-up to a reference value (e.g. 0.7),^19,20^ or used dose-response analyses,^21^ neither of which addressed the problem of finding thresholds associated with maximum effect on risk of events.

Use of arbitrary adherence thresholds with unknown clinical significance has important consequences. In research, it can result in misclassification bias (e.g. categorizing nonadherent patients as adherent), hinder analyses of pharmacologic and/or adherence improvement interventions, and introduce heterogeneity in the literature that undermines meta-analysis. In clinical practice, unvalidated adherence thresholds can give false confidence in the effectiveness of less-than-perfect adherence with consequent under-resourcing of efforts to improve it. It can also misguide reimbursement policies designed to encourage adequate adherence^45,55^ and lead to substantial direct and indirect costs due to preventable adverse outcomes caused by nonadherence. Perhaps most importantly, unvalidated adherence thresholds is likely to cause missed opportunities to identify and intervene in patients with poor adherence, likely resulting, in the case of AF, in preventable strokes and deaths.^3–8,10^

Our primary finding that the optimal threshold for DOAC for most outcomes is 0.9 has several implications. First, patients whose adherence is below this high level are at substantially greater risk of strokes. Second, this level of adherence is higher than conventionally considered “adherent”, meaning many patients may be mistaken in their belief that their 80-89% adherence level is sufficient to minimize risk of these devastating events. Such patients may need to be identified via interviews and/or prescription record screening and provided with education and/or adherence support focused on their own adherence barriers. Third, for quality of care and research purposes, if it is accepted that thresholds should be predictive of clinical outcomes,^14^ DOAC recipients should be classified as nonadherent when their PDC is <0.9. Such a high optimal threshold is consistent with evidence from other drugs for a range of conditions, including adalimumab and certolizumab for inflammatory bowel disease (86%),^51^ heart failure medications for event-free survival (88%),^56^ lipid-lowering therapies for all-cause mortality (84%),^50^ protease inhibitors (95%) and reverse transcriptase inhibitors (85-89%) for HIV viral suppression,^49^ and chronic hepatitis B antivirals for favorable virological outcome (90%).^47^

Our findings that the optimal threshold for VKA is 0.90 or 0.95 depending on the outcome and that the relationship between PDC and outcome risk is somewhat flat across the PDC spectrum also has several implications. First, the relationship between PDC and outcomes with VKA appears more complex than with DOAC. This may be because of the inherently complex pharmacodynamics of VKA with its narrow therapeutic index,^57^ longer duration of effect than DOACs,^58^ more variable anticoagulation effects at similar weight-normalized doses (possibly contributed to by pharmacogenetic heterogeneity),^59^ and unmeasured confounding due to differences between VKA and DOAC recipients. These causal explanations deserve empiric study. Time-in-therapeutic range (TTR) measurement obviates the importance of some of these variables but is not available for many screening programs or population-level research due to lack of laboratory data. However, our analyses based on a PDC measurement model validated for administrative data^31^ imply that the optimal threshold for VKA is much higher than conventionally thought, while at the same time being less clinically consequential than for DOACs. In other words, missing doses of VKA may be less consequential than missing DOAC doses. This is consistent with other recent findings.^10^

Our study is the first to derive OAC adherence thresholds that are rigorously optimized for clinical outcomes in AF patients. Prior attempts to identify optimal thresholds have critical methodological limitations. Kim et al.^20^ and Grymonprez et al.^19^ categorized PDC into arbitrary levels and inferred thresholds based on hazard ratios against a reference group, an approach that fails to statistically compare thresholds or evaluate model performance across them. Wirbka et al.^21^ employed a logistic growth model (ED80) but did not address the threshold optimization problem directly, instead reverting to a conventional 80%-of-effect paradigm. Notably, none of these studies systematically assessed whether model discrimination or fit peaked across adherence levels—a gap our biniLasso framework resolves by directly optimizing thresholds based on association with outcomes. As such, our results emerge from the most robust analytical approach yet applied to the important need for evidence-based adherence thresholds in OAC therapy.

Higher complexity models (continuous b-spline, linear) fit the data better than lower complexity models (2- or 1-threshold categorical), but the categorical models using the optimal adherence thresholds we derived still had excellent ability to classify patients in terms of their outcome risk, particularly for stroke outcomes. Despite this, it is valuable to know that the relationships are not linear and in fact, there appear to be portions of the PDC spectrum where outcome hazard strongly varies with changes in PDC, and other flat areas where it does not. There are also peaks and valleys for some outcomes, particularly with VKA. While it is vital to derive measures clinicians and patients can understand and take actions based on (e.g. categorical adherence thresholds), they will evidently obscure underlying complexity of the relationships between adherence and outcomes.

For several outcomes, despite the relationship being nonlinear, a linear model fit the data better than categorical threshold models. Clinicians clearly have the option to think and communicate linearly (e.g. for every 10% increase in DOAC adherence, risk of SSE decreases by 13%), but we believe that the optimal thresholds we derived are also statistically valid, were strongly associated with clinical outcomes in most cases, and potentially simpler for clinicians and patients to motivate adherence. Thresholds are also a good fit with the typical model of clinical decision-making where an action is taken or not taken depending on a particular variable or set of them. Furthermore, the thresholds we derived were significantly more valid for this purpose than the threshold of 0.8 commonly used.

Though the adherence goal for all chronic therapies is 100% to mimic the beneficial effects shown in randomized clinical trials, resources for supporting anticoagulated patients’ adherence are finite and trade-offs may be required. For example, our results suggest resources could more impactfully be devoted to helping DOAC recipients achieve >90% adherence than to, for example, screening VKA recipients’ prescription records to identify <80% adherence.

Furthermore, the nature and intensity of interventions to support adherence may need to be different if the goal is ≥90% adherence, compared to ≥80%. For warfarin, measuring INR remains the mainstay adherence monitoring, but for quality measurement and research programs using prescription fill data, it would be prudent to use PDC of 0.9 or 0.95 as the threshold for discriminating adherent from nonadherent VKA recipients.

Our findings for major bleeding deserve comment. Available evidence about the relationship between OAC adherence and major bleeding paints no clear picture, with some studies showing a positive association between PDC and major bleeding,^60–62^ some no association,^63–65^ and some a negative association.^10,66,67^ Our results are in the latter category, and are unintuitive if major bleeding is a dose-related toxicity.^68,69^ The reasons for this deserve further study via causal inference methods to help disentangle the true effects of adherence from confounder effects, particularly healthy user effects and unmeasured confounders.^70,71^ Nonetheless, the preponderance of available evidence supports the notion that efforts to achieve the efficacy gains of OAC adherence >90% need not be tempered by concerns about additional bleeding.

A key strength of our study is the robust datasets on which it was based, which are considered among the best available because of their complete coverage of the population due to BC’s single-payer system and mandatory capture of all prescription fills. Furthermore, our use of PDC and drug class at the time of event overcomes the prevalent limitation in the literature where PDC is averaged over long periods of follow-up, obscuring the critical few days/weeks leading up to an event since OACs are short-acting and confer no residual benefits after stopping.^72,73^ Furthermore, we have previously shown that long-term average PDC is of limited relevance for outcome studies due to its high variability in individuals.^32^ Although our data covered only through the end of 2019, this coincided with the beginning of the COVID-19 pandemic so our results are not confounded by disruptions to the medication supply system caused by the pandemic.

### Limitations

These results should be interpreted considering the study’s limitations. As with all research using PDC as an exposure, we assumed that filling a prescription correlates with consuming the medication, which may not always be true. For our multivariate modelling, some variables known to affect adherence (e.g., education level, adverse effects experienced, psychosocial factors) were absent from the administrative databases, limiting our ability to account for their contributions. Requiring at least two prescription fills for inclusion in the cohort (required for PDC calculation) meant that we excluded those who never initiated therapy or stopped after one prescription fill, a subpopulation with potentially different characteristics than those we studied. For example, patients experiencing serious OAC adverse effects immediately after starting would not be captured in our analyses, which likely primarily affected our major bleeding outcome. The available data did not include laboratory values, so we were unable to validate VKA adherence with INR values or include measures of renal function in study models. There were too few edoxaban recipients and too few of certain events (ICH, deaths due to bleeding, AF, or stroke) to permit analyses. The effects on outcome hazard we observed are associations and not necessarily causal in terms of PDC, despite adjustment for many covariates. For example, the differences between VKA and DOAC adherence thresholds we found may not be due to properties of the drugs themselves, but of the patients prescribed them. Although we included individualized stroke risk in our models (the distributions of which were known to be different between VKA and DOAC recipients), it is possible that other factors contributed to the differences. Though not essential for identifying clinically significant adherence thresholds as in the present analyses, causal inference methods could help in future investigations to identify the causes of the differences in adherence thresholds we found.

## CONCLUSIONS

For all studied outcomes, the optimal adherence thresholds for VKA and DOAC were roughly 0.95 and 0.9, respectively, and significantly higher than the conventional 0.8 threshold. The magnitude of outcome reduction for being above optimal thresholds for DOAC was much greater than for VKA, implying that the consequences of sub-threshold adherence are considerably more severe with DOACs than VKA. Our results highlight the need to consider updating the definition of nonadherence to PDC <0.9 for DOACs, and PDC<0.9 or <0.95 for VKA, depending on the outcome.

## Data Availability

Access to data provided by the Data Stewards is subject to approval but can be requested for research projects through the Data Stewards or their designated service providers. The following data sets were used in this study: Medical Services Plan (MSP), Discharge Abstract Database (DAD), Consolidation File, Vital Statistics Database, and PharmaNet. You can find further information regarding these data sets by visiting the PopData project webpage at: https://my.popdata.bc.ca/project_listings/17-149/collection_approval_dates. All inferences, opinions, and conclusions drawn in this publication are those of the author(s), and do not reflect the opinions or policies of the Data Steward(s).

## ACKNOWLEDGEMENTS

The authors are grateful to Dr. Anita I. Kapanen from University of British Columbia for her expert support of the data stewardship and ethical oversight of this study.

## Abbreviations

AF: atrial fibrillation
OAC: oral anticoagulant
PDC: proportion of days covered
VKA: vitamin K antagonist
DOAC: direct oral anticoagulant
SSE: stroke or systemic embolism
TIA: transient ischemic attack
REWarDS: Random Effects Warfarin Days Supply model
LASSO: least absolute shrinkage and selection operator
CGAM: Cox-based generalized additive model

## REFERENCES

1. Salmasi S, Loewen PS, Tandun R, Andrade JG, Vera MAD. Adherence to oral anticoagulants among patients with atrial fibrillation: a systematic review and meta-analysis of observational studies. BMJ Open. 2020;10:e034778.

2. Ozaki AF, Choi AS, Le QT, Ko DT, Han JK, Park SS, Jackevicius CA. Real-World Adherence and Persistence to Direct Oral Anticoagulants in Patients With Atrial Fibrillation. Circ: Cardiovasc Qual Outcomes. 2020;13:e005969.

3. Yao X, Abraham NS, Alexander GC, Crown W, Montori VM, Sangaralingham LR, Gersh BJ, Shah ND, Noseworthy PA. Effect of Adherence to Oral Anticoagulants on Risk of Stroke and Major Bleeding Among Patients With Atrial Fibrillation. J Am Hear Assoc. 2016;5:e003074.

4. Arbel A, Abu-Ful Z, Preis M, Cohen S, Saliba W. Adherence with direct oral anticoagulants in patients with atrial fibrillation: Trends, risk factors, and outcomes. J Arrhythmia. 2022;38:67–76.

5. Patsiou V, Samaras A, Kartas A, Moysidis DV, Papazoglou AS, Bekiaridou A, Baroutidou A, Ziakas A, Tzikas A, Giannakoulas G. Prognostic implications of adherence to oral anticoagulants among patients with atrial fibrillation: Insights from MISOAC-AF trial. J Cardiol. 2023;81:390– 396.

6. Alberts MJ, Peacock WF, Fields LE, Bunz TJ, Nguyen E, Milentijevic D, Schein JR, Coleman CI. Association between once-and twice-daily direct oral anticoagulant adherence in nonvalvular atrial fibrillation patients and rates of ischemic stroke. Int J Cardiol [Internet]. 2015;215:11–13. Available from: https://www.internationaljournalofcardiology.com/article/S0167-5273(16)30652-0/abstract

7. Borne RT, O’Donnell C, Turakhia MP, Varosy PD, Jackevicius CA, Marzec LN, Masoudi FA, Hess PL, Maddox TM, Ho PM. Adherence and outcomes to direct oral anticoagulants among patients with atrial fibrillation: findings from the veterans health administration. BMC Cardiovasc Disord. 2017;17:236–236.

8. Komen JJ, Heerdink ER, Klungel OH, Mantel-Teeuwisse AK, Forslund T, Wettermark B, Hjemdahl P. Long-term persistence and adherence with non-vitamin K oral anticoagulants in patients with atrial fibrillation and their associations with stroke risk. Eur Hear J - Cardiovasc Pharmacother. 2021;7:f72–f80.

9. Akagi Y, Iketaki A, Nakamura R, Yamamura S, Endo M, Morikawa K, Oikawa S, Ohta T, Tatsumi S, Suzuki T, Mizushima A, Koido K, Takahashi T. Association between Cerebral Infarction Risk and Medication Adherence in Atrial Fibrillation Patients Taking Direct Oral Anticoagulants. Healthcare. 2021;9:1313.

10. Safari A, Helisaz H, Salmasi S, Adelakun A, Vera MAD, Andrade JG, Deyell MW, Loewen P. Association Between Oral Anticoagulant Adherence and Serious Clinical Outcomes in Patients With Atrial Fibrillation: A Long-Term Retrospective Cohort Study. J Am Hear Assoc. 2024;13:e035639.

11. Shah KK, Touchette DR, Marrs JC. Research and scholarly methods: Measuring medication adherence. J Am Coll Clin Pharm. 2023;6:416–426.

12. Seabury SA, Dougherty JS, Sullivan J. Medication adherence as a measure of the quality of care provided by physicians. Am J Manag care. 2019;25:78–83.

13. Haynes RB, Taylor DW, Sackett DL, Gibson ES, Bernholz CD, Mukherjee J. Can simple clinical measurements detect patient noncompliance? Hypertension. 1980;2:757–64.

14. Baumgartner PC, Haynes RB, Hersberger KE, Arnet I. A Systematic Review of Medication Adherence Thresholds Dependent of Clinical Outcomes. Front Pharmacol. 2018;9:1290–1290.

15. Zielinski GD, Rein N van, Teichert M, Klok FA, Rosendaal FR, Meer FJM van der, Huisman MV, Cannegieter SC, Lijfering WM. Adherence to direct oral anticoagulant treatment for atrial fibrillation in the Netherlands: A surveillance study. Pharmacoepidemiol Drug Saf. 2021;30:1027–1036.

16. Sørensen R, Nielsen BJ, Pallisgaard JL, Lee CJ-Y, Torp-Pedersen C. Adherence with oral anticoagulation in non-valvular atrial fibrillation: a comparison of vitamin K antagonists and non-vitamin K antagonists. Eur Hear J Cardiovasc Pharmacother. 2017;3:151–156.

17. Salmasi S, Safari A, Vera MAD, Högg T, Lynd LD, Koehoorn M, Barry AR, Andrade JG, Deyell MW, Rush KL, Zhao Y, Loewen P. Adherence to direct or vitamin K antagonist oral anticoagulants in patients with atrial fibrillation: a long-term observational study. J Thromb Thrombolysis. 2024;57:437–444.

18. Farinha JM, Jones ID, Lip GYH. Optimizing adherence and persistence to non-vitamin K antagonist oral anticoagulant therapy in atrial fibrillation. Eur Hear J Suppl. 2022;24:A42–a55.

19. Grymonprez M, Steurbaut S, Capiau A, Vauterin D, Vaerenbergh FV, Mehuys E, Boussery K, Backer TLD, Lahousse L. Minimal Adherence Threshold to Non-Vitamin K Antagonist Oral Anticoagulants in Patients with Atrial Fibrillation to Reduce the Risk of Thromboembolism and Death: A Nationwide Cohort Study. Cardiovasc Drugs Ther. 2025;39:107–117.

20. Kim D, Yang P-S, Jang E, Yu HT, Kim T-H, Uhm J-S, Kim J-Y, Sung J-H, Pak H-N, Lee M-H, Lip GYH, Joung B. The optimal drug adherence to maximize the efficacy and safety of non-vitamin K antagonist oral anticoagulant in real-world atrial fibrillation patients. EP Eur. 2019;22:547–557.

21. Wirbka L, Haefeli WE, Meid AD, Group AS. Estimated Thresholds of Minimum Necessary Adherence for Effective Treatment with Direct Oral Anticoagulants – A Retrospective Cohort Study in Health Insurance Claims Data. Patient preference adherence. 2021;15:2209–2220.

22. Stauffer ME, Hutson P, Kaufman AS, Morrison A. The Adherence Rate Threshold is Drug Specific. Drugs R D [Internet]. 2017;17:645–653. Available from: https://pubmed.ncbi.nlm.nih.gov/29076037

23. Ark T, Kesselring S, Hills B, McGrail K. Population Data BC: Supporting population data science in British Columbia. Int J Popul Data Sci. 2019;4:1133.

24. Elm E von, Altman DG, Egger M, Pocock SJ, Gøtzsche PC, Vandenbroucke JP. The Strengthening the Reporting of Observational Studies in Epidemiology (STROBE) Statement: Guidelines for Reporting Observational Studies. Plos Med. 2007;4:e296.

25. Benchimol EI, Smeeth L, Guttmann A, Harron K, Moher D, Petersen I, Sørensen HT, Elm E von, Langan SM, Committee RW. The REporting of studies Conducted using Observational Routinely-collected health Data (RECORD) Statement. Plos Med. 2015;12:e1001885.

26. Langan SM, Schmidt SA, Wing K, Ehrenstein V, Nicholls SG, Filion KB, Klungel O, Petersen I, Sorensen HT, Dixon WG, Guttmann A, Harron K, Hemkens LG, Moher D, Schneeweiss S, Smeeth L, Sturkenboom M, Elm E von, Wang SV, Benchimol EI. The reporting of studies conducted using observational routinely collected health data statement for pharmacoepidemiology (RECORD-PE). Bmj. 2018;363:k3532.

27. Suissa S. Immortal time bias in observational studies of drug effects. Pharmacoepidemiol Drug Saf. 2007;16:241–249.

28. Dalli LL, Kilkenny MF, Arnet I, Sanfilippo FM, Cummings DM, Kapral MK, Kim J, Cameron J, Yap KY, Greenland M, Cadilhac DA. Towards better reporting of the proportion of days covered method in cardiovascular medication adherence: A scoping review and new tool TEN-SPIDERS. Br J Clin Pharmacol. 2022;88:4427–4442.

29. Karve S, Cleves MA, Helm M, Hudson TJ, West DS, Martin BC. Prospective validation of eight different adherence measures for use with administrative claims data among patients with schizophrenia. Value Heal. 2009;12:989–95.

30. Prieto-Merino D, Mulick A, Armstrong C, Hoult H, Fawcett S, Eliasson L, Clifford S. Estimating proportion of days covered (PDC) using real-world online medicine suppliers’ datasets. J Pharm Polic Pr. 2021;14:113.

31. Salmasi S, Högg T, Safari A, Vera MAD, Lynd LD, Koehoorn M, Barry AR, Andrade JG, Loewen P. The Random Effects Warfarin Days’ Supply (REWarDS) model: Development and validation of a novel method to estimate exposure to warfarin using administrative data. Am J Epidemiol. 2022;191:1116–1124.

32. Salmasi S, Vera MAD, Safari A, Lynd LD, Koehoorn M, Barry AR, Andrade JG, Deyell MW, Rush K, Zhao Y, Loewen P. Longitudinal Oral Anticoagulant Adherence Trajectories in Patients With Atrial Fibrillation. J Am Coll Cardiol. 2021;78:2395–2404.

33. Lip GY, Nieuwlaat R, Pisters R, Lane DA, Crijns HJ. Refining clinical risk stratification for predicting stroke and thromboembolism in atrial fibrillation using a novel risk factor-based approach: the euro heart survey on atrial fibrillation. Chest. 2010;137:263–72.

34. Lip GYH, Frison L, Halperin JL, Lane DA. Comparative Validation of a Novel Risk Score for Predicting Bleeding Risk in Anticoagulated Patients With Atrial Fibrillation The HAS-BLED (Hypertension, Abnormal Renal/Liver Function, Stroke, Bleeding History or Predisposition, Labile INR, Elderly, Drugs/Alcohol Concomitantly) Score. J Am Coll Cardiol. 2011;57:173– 180.

35. Schaefer JK, Errickson J, Li Y, Kong X, Alexandris-Souphis T, Ali MA, Decamillo D, Haymart B, Kaatz S, Kline-Rogers E, Kozlowski JH, Krol GD, Shankar SR, Sood SL, Froehlich JB, Barnes GD. Adverse Events Associated With the Addition of Aspirin to Direct Oral Anticoagulant Therapy Without a Clear Indication. JAMA Intern Med. 2021;181:817–824.

36. Quan H, Sundararajan V, Halfon P, Fong A, Burnand B, Luthi JC, Saunders LD, Beck CA, Feasby TE, Ghali WA. Coding algorithms for defining comorbidities in ICD-9-CM and ICD-10 administrative data. Méd Care. 2005;43:1130–9.

37. Sharma N, Schwendimann R, Endrich O, Ausserhofer D, Simon M. Comparing Charlson and Elixhauser comorbidity indices with different weightings to predict in-hospital mortality: an analysis of national inpatient data. BMC Heal Serv Res. 2021;21:13.

38. Masnoon N, Shakib S, Kalisch-Ellett L, Caughey GE. What is polypharmacy? A systematic review of definitions. Bmc Geriatr. 2017;17:230.

39. Buuren S van, Groothuis-Oudshoorn K. mice : Multivariate Imputation by Chained Equations in R. J Stat Softw. 2011;45.

40. Safari A, Helisaz H, Loewen P. biniLasso: Automated cut-point detection via sparse cumulative binarization. *arxiv*.org [Internet]. 2025;1–10. Available from: https://arxiv.org/abs/2503.16687

41. Simon N, Friedman J, Hastie T, Tibshirani R. Regularization Paths for Cox’s Proportional Hazards Model via Coordinate Descent. J Stat Softw. 2011;39:1–13.

42. Vittinghoff E, McCulloch CE. Relaxing the rule of ten events per variable in logistic and Cox regression. Am J Epidemiol. 2007;165:710–8.

43. Holm S. A Simple Sequentially Rejective Multiple Test Procedure. Scandinavian Journal of Statistics [Internet]. 1979;6:65–70. Available from: https://www.jstor.org/stable/4615733

44. Hastie TJ, Tibshirani RJ. Generalized Additive Models. 2017;82–104.

45. Gellad WF, Thorpe CT, Steiner JF, Voils CI. The myths of medication adherence. Pharmacoepidemiol Drug Saf. 2017;26:1437–1441.

46. Andrade SE, Kahler KH, Frech F, Chan KA. Methods for evaluation of medication adherence and persistence using automated databases. Pharmacoepidemiol Drug Saf. 2006;15:565–74; discussion 575-7.

47. Allard NL, MacLachlan JH, Dev A, Dwyer J, Srivatsa G, Spelman T, Thompson AJ, Cowie BC. Adherence in chronic hepatitis B: associations between medication possession ratio and adverse viral outcomes. BMC Gastroenterol [Internet]. 2020;20:140. Available from: http://ovidsp.ovid.com/ovidweb.cgi?T=JS&PAGE=reference&D=med18&NEWS=N&AN=32381025

48. Izano MA, Neugebauer R, Ettinger B, Hui R, Chandra M, Adams AL, Niu F, Ott SM, Lo JC. Using Pharmacy Data and Adherence to Define Long-Term Bisphosphonate Exposure in Women. J Manag Care Spéc Pharm [Internet]. 2019;25:719–723. Available from: http://ovidsp.ovid.com/ovidweb.cgi?T=JS&PAGE=reference&D=med16&NEWS=N&AN=31134854

49. Viswanathan S, Justice AC, Alexander GC, Brown TT, Gandhi NR, McNicholl IR, Rimland D, Rodriguez-Barradas MC, Jacobson LP. Adherence and HIV RNA Suppression in the Current Era of Highly Active Antiretroviral Therapy. Jaids J Acquir Immune Defic Syndromes. 2015;69:493–498.

50. Zongo A, Simpson S, Johnson JA, Eurich DT. Optimal threshold of adherence to lipid lowering drugs in predicting acute coronary syndrome, stroke, or mortality: A cohort study. PLoS ONE. 2019;14:e0223062.

51. Govani SM, Noureldin M, Higgins PDR, Heisler M, Saini SD, Stidham RW, Waljee JF, Waljee AK. Defining an Optimal Adherence Threshold for Patients Taking Subcutaneous Anti-TNFs for Inflammatory Bowel Diseases. Am J Gastroenterol. 2018;113:276–282.

52. Burnier M. Is There a Threshold for Medication Adherence? Lessons Learnt From Electronic Monitoring of Drug Adherence. Front Pharmacol [Internet]. 2019;9:1540–1540. Available from: https://pubmed.ncbi.nlm.nih.gov/30687099

53. Yaegashi H, Kirino S, Remington G, Misawa F, Takeuchi H. Adherence to Oral Antipsychotics Measured by Electronic Adherence Monitoring in Schizophrenia: A Systematic Review and Meta-analysis. CNS Drugs. 2020;34:579–598.

54. Alliance PQ. PQA Adherence Measures [Internet]. [cited 2024 Oct 11];Available from: https://www.pqaalliance.org/adherence-measures

55. Parekh N, Munshi KD, Hernandez I, Gellad WF, Henderson R, Shrank WH. Impact of Star Rating Medication Adherence Measures on Adherence for Targeted and Nontargeted Medications. Value Heal. 2019;22:1266–1274.

56. Wu JR, Moser DK, Jong MJD, Rayens MK, Chung ML, Riegel B, Lennie TA. Defining an evidence-based cutpoint for medication adherence in heart failure. Am Hear J. 2009;157:285–91.

57. Holford NHG. Clinical Pharmacokinetics and Pharmacodynamics of Warfarin. Clin Pharmacokinet. 1986;11:483–504.

58. Eikelboom J, Merli G. Bleeding with direct oral anticoagulants vs warfarin: clinical experience. Am J Emerg Med. 2016;34:3–8.

59. Fahmi AM, Elewa H, Jilany IE. Warfarin dosing strategies evolution and its progress in the era of precision medicine, a narrative review. Int J Clin Pharm. 2022;44:599–607.

60. Kang H-R, Jones BL, Lo-Ciganic W-H, DeRemer CE, Dietrich EA, Huang P-L, Park H. Trajectories of adherence to extended treatment with warfarin and risks of recurrent venous thromboembolism and major bleeding. Res Pr Thromb Haemost. 2023;7:100131.

61. An J, Bider Z, Luong TQ, Cheetham TC, Lang DT, Fischer H, Reynolds K. Long-Term Medication Adherence Trajectories to Direct Oral Anticoagulants and Clinical Outcomes in Patients With Atrial Fibrillation. J Am Hear Assoc. 2021;10:e021601.

62. Lee WK, Woo SI, Hyun DK, Jung S-Y, Kim M-S, Lee J. Impact of treatment adherence on the effectiveness and safety of oral anticoagulants in patients with atrial fibrillation: a retrospective cohort study. European Hear J - Qual Care Clin Outcomes. 2022;9:216–226.

63. Kang H-R, Jones BL, Lo-Ciganic W-H, DeRemer CE, Dietrich EA, Huang P-L, Wilson DL, Park H. Trajectories of adherence to extended treatment with direct oral anticoagulants and risks of recurrent venous thromboembolism and major bleeding. J Manag Care Spéc Pharm. 2023;29:1219–1230.

64. McHorney CA, Peterson ED, Ashton V, Laliberté F, Crivera C, Germain G, Sheikh N, Schein J, Lefebvre P. Modeling the impact of real-world adherence to once-daily (QD) versus twice-daily (BID) non-vitamin K antagonist oral anticoagulants on stroke and major bleeding events among non-valvular atrial fibrillation patients. Curr Méd Res Opin. 2019;35:653–660.

65. Hurtado-Navarro I, García-Sempere A, Rodríguez-Bernal C, Santa-Ana-Tellez Y, Peiró S, Sanfélix-Gimeno G. Estimating Adherence Based on Prescription or Dispensation Information: Impact on Thresholds and Outcomes. A Real-World Study With Atrial Fibrillation Patients Treated With Oral Anticoagulants in Spain. Front Pharmacol. 2018;9:1353.

66. Akao M, Yamashita T, Atarashi H, Ikeda T, Koretsune Y, Okumura K, Shimizu W, Suzuki S, Tsutsui H, Toyoda K, Hirayama A, Yasaka M, Yamaguchi T, Teramukai S, Kimura T, Morishima Y, Takita A, Inoue H. Comprehension of Nonvalvular Atrial Fibrillation and Anticoagulant Adherence in Elderly Patients in a Subcohort Study of the All Nippon Atrial Fibrillation in the Elderly Registry. Am J Cardiol. 2023;204:159–167.

67. Millenaar D, Schumacher H, Brueckmann M, Eikelboom JW, Ezekowitz M, Slawik J, Ewen S, Ukena C, Wallentin L, Connolly S, Yusuf S, Böhm M. Cardiovascular Outcomes According to Polypharmacy and Drug Adherence in Patients with Atrial Fibrillation on Long-Term Anticoagulation (from the RE-LY Trial). Am J Cardiol. 2021;149:27–35.

68. Giugliano RP, Ruff CT, Braunwald E, Murphy SA, Wiviott SD, Halperin JL, Waldo AL, Ezekowitz MD, Weitz JI, Špinar J, Ruzyllo W, Ruda M, Koretsune Y, Betcher J, Shi M, Grip LT, Patel SP, Patel I, Hanyok JJ, Mercuri M, Antman EM. Edoxaban versus warfarin in patients with atrial fibrillation. N Engl J Med. 2013;369:2093–104.

69. Hylek EM, Go AS, Chang Y, Jensvold NG, Henault LE, Selby JV, Singer DE. Effect of Intensity of Oral Anticoagulation on Stroke Severity and Mortality in Atrial Fibrillation. N Engl J Med. 2003;349:1019–1026.

70. Shrank WH, Patrick AR, Brookhart MA. Healthy User and Related Biases in Observational Studies of Preventive Interventions: A Primer for Physicians. J Gen Intern Med. 2011;26:546– 550.

71. Robins JM, Hernán MÁ, Brumback B. Marginal Structural Models and Causal Inference in Epidemiology. Epidemiology. 2000;11:550–560.

72. Kearon C, Kahn SR. Long-term treatment of venous thromboembolism. Blood. 2020;135:317–325.

73. Julia S, James U. Direct Oral Anticoagulants: A Quick Guide. Eur Cardiol [Internet]. 2017;12:40–45. Available from: https://pubmed.ncbi.nlm.nih.gov/30416551

